# Leveraging informative missing data to learn about acute respiratory distress syndrome and mortality in long-term hospitalized COVID-19 patients throughout the years of the pandemic

**DOI:** 10.1101/2023.12.18.23300181

**Authors:** Emily Getzen, Amelia LM Tan, Gabriel Brat, Gilbert S. Omenn, Zachary Strasser, The Consortium for Clinical Characterization of COVID-19 by EHR (4CE) (Collaborative Group/Consortium), Qi Long, John H. Holmes, Danielle Mowery

**Author notes:** co-first authors (Emily Getzen, Amelia LM Tan). co-senior authors (Qi Long, John H. Holmes, Danielle Mowery).

## Abstract

Electronic health records (EHRs) contain a wealth of information that can be used to further precision health. One particular data element in EHRs that is not only under-utilized but oftentimes unaccounted for is missing data. However, missingness can provide valuable information about comorbidities and best practices for monitoring patients, which could save lives and reduce burden on the healthcare system. We characterize patterns of missing data in laboratory measurements collected at the University of Pennsylvania Hospital System from long-term COVID-19 patients and focus on the changes in these patterns between 2020 and 2021. We investigate how these patterns are associated with comorbidities such as acute respiratory distress syndrome (ARDS), and 90-day mortality in ARDS patients. This work displays how knowledge and experience can change the way clinicians and hospitals manage a novel disease. It can also provide insight into best practices when it comes to patient monitoring to improve outcomes.

## Introduction

Traditionally, electronic health records (EHRs) were utilized in health systems for supporting billing purposes and healthcare operations. Today, secondary use of EHR data is leveraged to better predict adverse health outcomes, map disease trajectories, and evaluate treatment efficacy. More recently, they have been used to investigate the novel Coronavirus Disease (COVID-19) caused by the SARS-CoV-2 virus. However, there are many existing challenges that come with the analysis of EHRs– they require a great deal of pre-processing to generate a dataset with well-defined features of a patient’s clinical case ^1^. They do not have standardized formatting^1^, can introduce collection bias, and contain diagnostic coding errors^2,3^. There is also a lack of interoperability between EHR systems^3,4^, and some institutions / geographic regions may not have the technological or financial resources to implement EHRs^5^. It follows that the most frequently reported barrier to EHRs usability is missing data, or data that would have existed if measured, but was not measured^4, 6-9^.

Missing data is not always necessarily a reflection of quality of care from an informatics perspective^10^. In hospitalized patients, for example, laboratory measurements are taken at a certain frequency. This frequency might vary depending on what a patients’ condition is, or how severe they are. The underlying lab values always exist at any given time point, but whether or not they are *measured* or observed is subject to change. Thus, we might expect hospitalized patients with less severe conditions to have more missing data compared to patients with more severe conditions, as a result of the latter being more closely monitored with measurements taken at a higher frequency.

There are a variety of ways that missingness can be informative. Groups of measurements that tend to always be missing and present together can denote a panel used to monitor for specific diseases^10^. Measurements that tend not to be associated under normal circumstances but sync up later in the course of admission can indicate the emergence of a condition– for example, when LDH and Fibrinogen are ordered together, it can indicate suspicion of infection^10^. Furthermore, these patterns could be used to identify potential comorbidities, and help us learn about the clinical trajectory of a hospitalized patient. There are a variety of previous works that leverage informative missing data to improve prediction models in medical settings. Groenwold et al., 2020 determined that informative missingness can be incorporated into a clinical prediction model using EHRs^11^. Singh et al., 2021 evaluated the effectiveness of missingness features on machine learning models specifically for tasks like length of stay prediction.^12^ Informative missingness has additionally been investigated in longitudinal cohort studies^13-14^, genetic association studies and genotype analysis^15-19^, and case/control studies^20^.

Over the course of the pandemic, physicians learned how to better manage the disease, thus changes in trends from 2020 to 2021 might indicate emerging subphenotypes or better laboratory monitoring practices^21-23^. Information about comorbidities and best practices for monitoring patients is valuable for saving lives and reducing burden on the healthcare system. Thus, in this work, we leverage informative missingness across the years of the pandemic to learn more about comorbidities in long-term patients such as acute respiratory distress syndrome, and use it to predict adverse outcomes such as 90-day mortality in ARDS patients. We focus specifically on acute respiratory distress (ARDS) as a comorbidity, as it is a dangerous and potentially fatal respiratory condition for hospitalized COVID-19 patients.

## Methods

This retrospective observational study of EHRs was reviewed and approved by the ethics and institutional review boards at the University of Pennsylvania. For this study, we analyzed the EHR data from the University of Pennsylvania (UPenn) site via the resources of the Consortium for the Clinical Characterization of COVID-19 by EHR (4CE). The UPenn site data was generated from five hospitals with 2,469 beds, 118,188 inpatient discharges per year^10^. The inclusion criterion for long-term hospitalized COVID-19 patients include a positive COVID-19 polymerase chain reaction test on or during admission to the inpatient setting, and the patient had to have stayed in the hospital for at least 14 days. For each patient, we restrict data to their first hospital admission. We focus our analyses on 16 laboratory tests measured over the entire admission, which are selected because their abnormal values have been associated with poor outcomes in COVID-19 patients^24^.

**Table 1.**
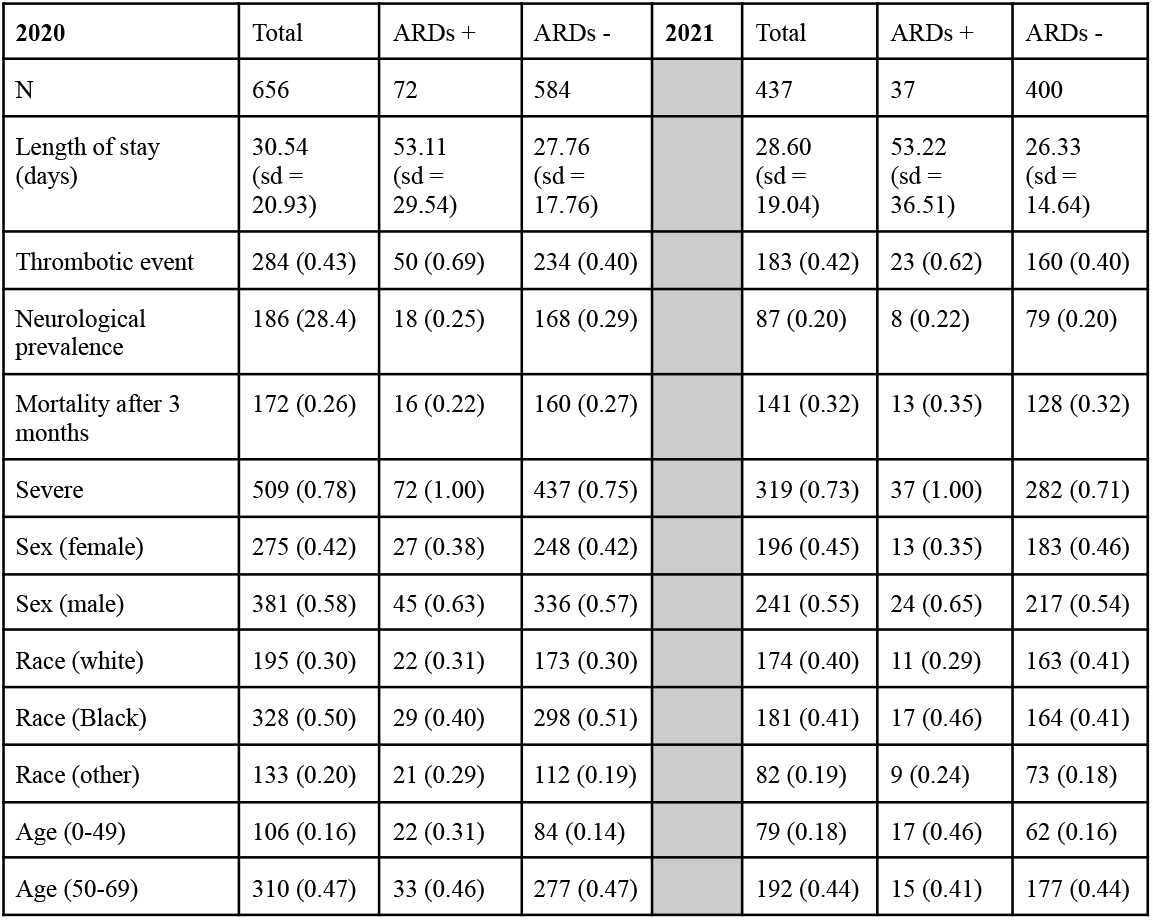
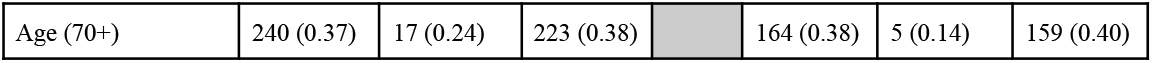
Characteristics of long-term patients (patients in hospital > 14 days)

| **2020** | Total | ARDs + | ARDs - | **2021** | Total | ARDs + | ARDs - |
| --- | --- | --- | --- | --- | --- | --- | --- |
| N | 656 | 72 | 584 |  | 437 | 37 | 400 |
| Length of stay (days) | 30.54 (sd = 20.93) | 53.11 (sd = 29.54) | 27.76 (sd = 17.76) |  | 28.60 (sd = 19.04) | 53.22 (sd = 36.51) | 26.33 (sd = 14.64) |
| Thrombotic event | 284 (0.43) | 50 (0.69) | 234 (0.40) |  | 183 (0.42) | 23 (0.62) | 160 (0.40) |
| Neurological prevalence | 186 (28.4) | 18 (0.25) | 168 (0.29) |  | 87 (0.20) | 8 (0.22) | 79 (0.20) |
| Mortality after 3 months | 172 (0.26) | 16 (0.22) | 160 (0.27) |  | 141 (0.32) | 13 (0.35) | 128 (0.32) |
| Severe | 509 (0.78) | 72 (1.00) | 437 (0.75) |  | 319 (0.73) | 37 (1.00) | 282 (0.71) |
| Sex (female) | 275 (0.42) | 27 (0.38) | 248 (0.42) |  | 196 (0.45) | 13 (0.35) | 183 (0.46) |
| Sex (male) | 381 (0.58) | 45 (0.63) | 336 (0.57) |  | 241 (0.55) | 24 (0.65) | 217 (0.54) |
| Race (white) | 195 (0.30) | 22 (0.31) | 173 (0.30) |  | 174 (0.40) | 11 (0.29) | 163 (0.41) |
| Race (Black) | 328 (0.50) | 29 (0.40) | 298 (0.51) |  | 181 (0.41) | 17 (0.46) | 164 (0.41) |
| Race (other) | 133 (0.20) | 21 (0.29) | 112 (0.19) |  | 82 (0.19) | 9 (0.24) | 73 (0.18) |
| Age (0-49) | 106 (0.16) | 22 (0.31) | 84 (0.14) |  | 79 (0.18) | 17 (0.46) | 62 (0.16) |
| Age (50-69) | 310 (0.47) | 33 (0.46) | 277 (0.47) |  | 192 (0.44) | 15 (0.41) | 177 (0.44) |
| Age (70+) | 240 (0.37) | 17 (0.24) | 223 (0.38) |  | 164 (0.38) | 5 (0.14) | 159 (0.40) |

Of note, because we are restricting our cohort to patients who had at least 14 days worth of data in their hospital admission, we may have a smaller population of patients 70+ who are ARDS positive due to the fact that these patients are less likely to survive as long with the condition.

In this study, we define missingness as *the absence of a lab measurement during a particular time point; in this case, within a day*^10^. Thus, throughout a patients’ course of admission, the proportion of missing values for a particular lab would be the proportion of days that the lab was not measured. Again, it is important to note that in this setting, missing data does not necessarily reflect quality of clinical care delivered. It could be routine for a certain lab to be measured once every three days, and that would be reflected as 66.7% missing data for a particular patient.

We define two “phases” of COVID-19, or time periods in which we observe patients. Phase 1 is defined as the time period between January 1 2020 to December 31 2020 and Phase 2 is defined as the time period between January 1 2021 to December 31 2021. We also restrict hospital admissions to the first 14 days; thus, each patient has the same denominator when calculating the proportion of missing data for a given lab.

### Identifying biologically meaningful lab clusters via LDA topic modeling

We first characterized the relationships between the labs based on their missingness patterns with the goal of clustering labs with similar missingness patterns. This allows us to reduce our feature space and the number of statistical tests. To identify such patterns, we employed Latent Dirichlet Allocation (LDA) topic modeling to identify labs that tend to be missing / present together at the different phases of the pandemic.

LDA topic modeling is a generative probabilistic approach applied to model collections of discrete data, which in practice is typically text corpora^25^. Each topic is characterized by a distribution over the words in the documents, and documents are represented as random mixtures over latent topics. Using the same approach as Tan et al., 2023; the “words” are defined as the individual labs, and each “document” is defined as the sum of missing indicators (whether or not a lab was missing on a particular day) for each lab across the specified time period. The “topics” are the groups / clusters of similar labs based on those patterns. In essence, we are identifying which labs tend to be missing together. For more information on LDA topic modeling, see Blei et al., 2003.

Similar to Tan et al., 2023; we also use four metrics to determine the optimal number of topics to learn from the data. In this work, we evaluate a range of three to seven topics. We assess which of these maximized: *held-out likelihood, semantic coherence*, and *lower bound on the marginal likelihood*. The held-out likelihood provides a measure of how predictive the model is on unseen documents, and the semantic coherence measures the tendency of a topic’s high probability words to co-occur in the same document. We also assess which number of topics minimized the residuals. In the LDA model, every laboratory test has a non-zero probability of belonging to a given topic, with some labs having a higher probability than others.

We specifically assess the lab-topic probabilities to characterize the relationships of the labs to one another– if two labs have a high probability of belonging to topic A, and low probabilities of belonging to topics B and C, these labs likely tend to be missing together / ordered together more often. If LDA topic modeling generates two topics in which 70% of the labs that make up the 95% probability mass are the same, we simply average the two topics.

To obtain the smaller cluster of labs that mostly characterize a topic, a lab has to make up more than 10% of the probability mass, and the labs that satisfy this criterion must make up more than 70% of the probability mass. Based on this criterion, if a lab appears in two topics or does not appear in any topic, that lab will not be assigned to a cluster and analyzed separately. This is also the case if a lab is assigned to a cluster in 2020, but not 2021.

### Associating lab missingness clusters to Acute Respiratory Distress Syndrome

We were interested to see if missing data in the laboratory measurements was associated in some way to ARDS, and what that might say about the clinical trajectory of a patient. Based on the LDA topic modeling results, we calculate missingness scores for each cluster and each standalone lab (further referred to overall as lab features), which is the average proportion of missing data for each lab feature. Thus, at a specified time, each patient has X missing data scores for each lab feature, which is essentially the average proportion of missing data.

ARDS was defined by the presence of two diagnosis codes 518 (ICD-9) and J80 (ICD-10). To strengthen the validity of the relationship between missing data that we identified, we incorporated methods from causal inference in our models. We first convert each lab feature’s missingness scores to a binary variable that denotes which patients have ‘more missing data’ in a particular lab feature (> median). We build propensity score models via logistic regression with ‘more missing data in lab feature X’ as our treatment variable, and demographic / health status variables as covariates. Demographic variables include race, age, and sex; while health status variables include the presence of a thrombotic event, neurological event, and length of stay (ICD codes corresponding to thrombotic and neurological events in the supplement). We then used inverse probability weighting (IPW) on the propensity scores in logistic regression models to associate the continuous missingness scores for each lab feature with the presence of ARDS. Thus, our models are robust to potential confounding by race, age, sex, and health status. We control for false discovery rate (FDR) when determining significance at alpha = 0.05 for a single test.

### Leveraging lab missingness features in ARDS patients to predict 90-day mortality

Finally, we wanted to know whether or not we could further identify best practices to maximize better clinical outcomes in patients with comorbidities such as ARDS. Thus, we restricted our cohort to only patients with ARDS, and denoted a patient as positive for 90-day mortality if their death date occurred within three months after their 14-day observation period. We use a very similar approach to above and use IPW of propensity scores to account for potential confounding by demographic and health-status variables.

## Results

### Feature selection via LDA topic modeling of missing data in laboratory measurements

We employed LDA topic modeling on the counts of missing values in each lab for each long-term patient for the first 14 days. This led to two different topic models– 2020 and 2021.

**Figure 1.**
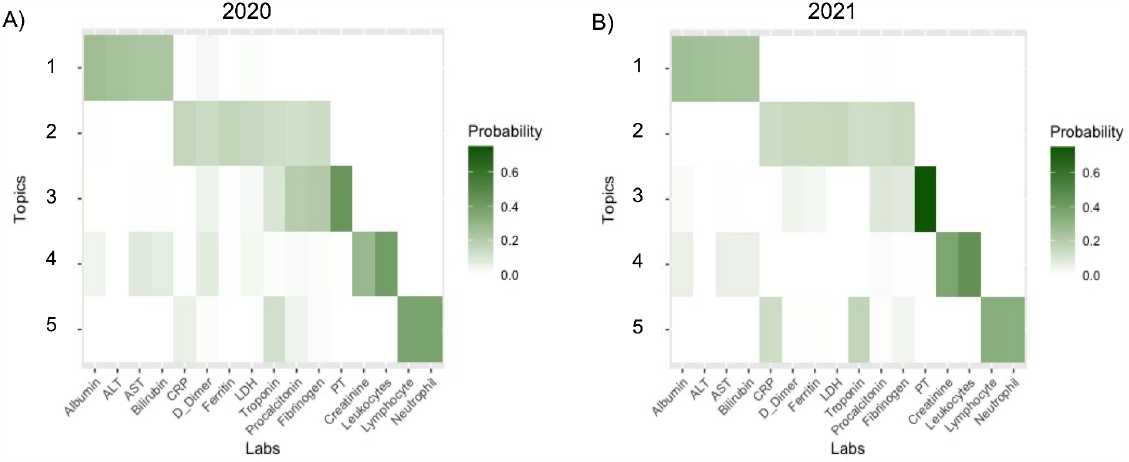
Topic modeling results over phase (2020, 2021). Darker colors indicate a higher probability of a lab belonging to a particular topic.

Labs with darker colors in a particular topic indicate that the lab contributes more to the probability mass of the topic. Two topics that contribute more probability mass than other labs are more likely to be associated (present / missing together). Based on the criterion defined in the Methods Section for assigning labs in a topic to a cluster, we identified four clusters which are similar to the clusters identified in Tan et al., 2023 at multiple hospital sites. Thus, we label them similarly based on biological implication:

1. Acute Inflammation: Ferritin, Lactate Dehydrogenase (LDH), D-Dimer, C-reactive Protein (CRP)
2. Infection-only: Lymphocytes, Neutrophils
3. Renal/Infection: Leukocytes, Creatinine
4. Liver: Albumin, Aspartate Aminotransferase (AST), Alanine Aminotransferase (ALT), Bilirubin

We also identified four stand-alone labs:

1. Prothrombin (PT)
2. Fibrinogen
3. Procalcitonin
4. Troponin

Most likely, the reason that Leukocytes did not appear in a cluster with Lymphocytes / Neutrophils is due to the fact that this site does complete blood counts without the differential, in which case leukocytes would be tested without lymphocytes / neutrophils. The association between leukocytes and creatinine may have to do with clinicians looking for kidney injury due to infection.

### Associations of lab-missingness features with ARDS

**Figure 2.**
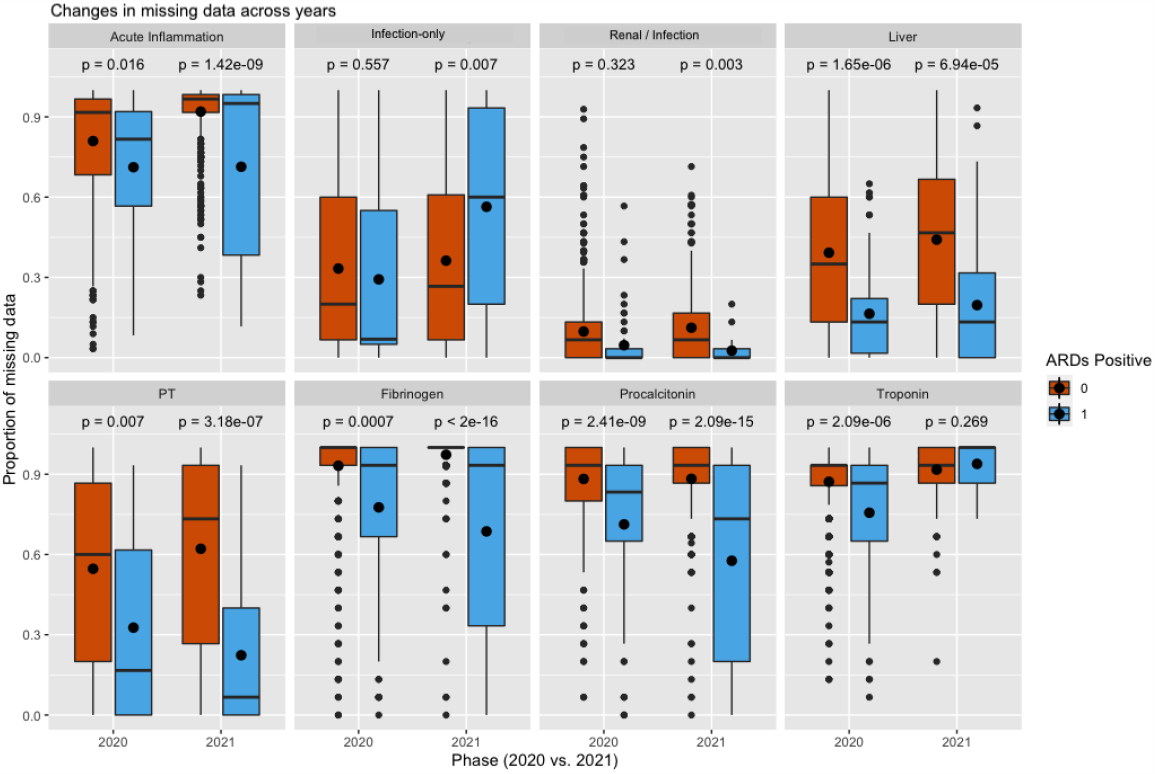
Comparison of the proportion of missing data in each lab feature across all patients that have ARDS versus those that do not

To identify relationships between missing data in certain lab groups and ARDS, we built univariate logistic regression models that associate each lab group’s missingness scores with whether or not a patient is positive for ARDS. Our models use inverse probability weighting of propensity scores that quantify how likely a patient is to have more missing data based demographics and health-status. Thus, our findings are more robust to potential confounding.

In 2020, having ARDS was associated with less missingness in all lab features except for the infection and renal labs. In 2021, missingness in the renal/infection labs goes from being non-significant to significant, and troponin goes in the other direction. We also observe that in 2021, *more* missingness in the infection-only labs is associated with having ARDS.

### Associations of lab-missingness features in ARDS patients and mortality outcomes

**Figure 3.**
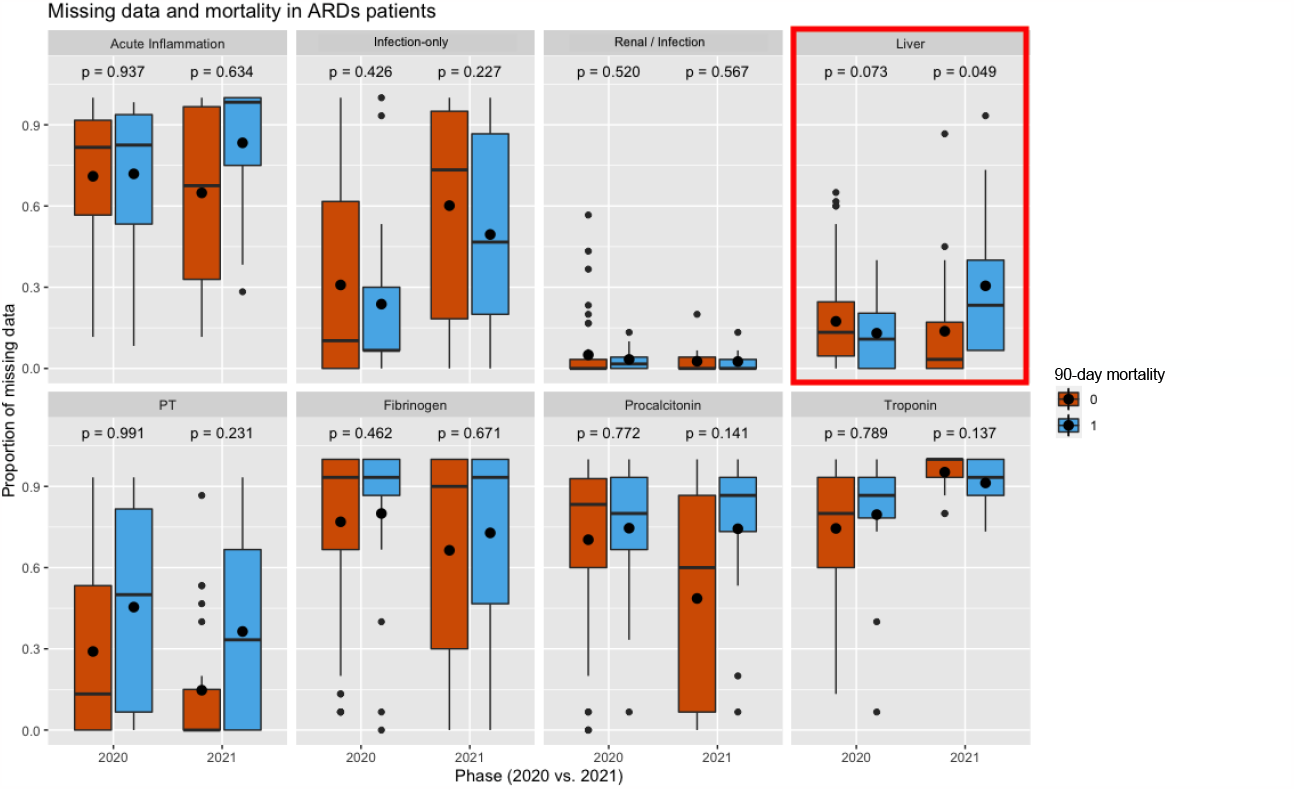
Comparison of the proportion of missing data in each lab feature across ARDS patients that experience 90-day mortality vs. those that do not.

To identify relationships between missing data in ARDS patients and 90-day mortality, we again built univariate logistic regression models that associate each lab group’s missingess scores with whether or not a patient passed away within three months after the observation period. Similar to above, our models use IPW of propensity scores to ensure robustness to demographic and health status variables.

In 2020, we do not observe any strong trends in which missingness is associated with 90-day mortality for ARDS patients. However, in 2021, more missingness in the liver labs is nearly associated with 90-day mortality, which could indicate the importance of monitoring ARDS patients’ liver function to potentially minimize mortality.

Although we observe that patients with more missingness in PT and Procalcitonin in 2021 experience 90-day mortality at a higher rate, this is not as significant. Since the p-values are quite high despite a large effect size, it is possible that there is potential confounding. After some investigation, we found that a much higher proportion of patients who survived had a thrombotic event (79% versus 38% in 2020, 75% versus 38% in 2021). Upon investigating the ages of these patients, we also found that patients with ARDS *and* a thrombotic event tend to be younger– in 2020 only 16% of ARDS patients with a thrombotic event were over the age of 70, whereas 41% of ARDS patients *without* a thrombotic event were over the age of 70. In 2021, these percentages are 4% and 29%, respectively. Thus, the confounding likely stems from age and selection bias likely due to older patients with ARDS *and* thrombotic events not surviving the 14 day observation period.

## Discussion

In this work, we assess how missing data in long-term hospitalized COVID-19 patients relates to patient clinical outcomes such as ARDS and within ARDS patients, how it predicts 90-day mortality. We leverage LDA topic modeling to reduce our feature space and group laboratory measurements into biologically meaningful clusters. We identified four clusters that were consistent in 2020 as well as 2021: acute inflammation labs, renal labs, infection-only labs, and liver labs. These clusters match 3 of the 4 biologically meaningful clusters identified in Tan et al., 2023. We were left with four standalone labs: PT, fibrinogen, procalcitonin, and troponin. We found that in 2020 less missing data in all of the lab features besides renal/infection and infection-only was associated with patients having ARDS. In 2021, however, this relationship became significant for the renal/infection labs, but not significant for troponin. In addition, *more* missingness in the infection-only labs was associated with patients having ARDS. We found that in 2021, more missing data in the liver labs was nearly predictive of 90-day mortality. Although less significant, we also observed that patients who experienced 90-day mortality had more missingness in PT and procalcitonin– this is likely due to selection bias in the ages of patients who survived the 14-day observation period with ARDS *and* thrombotic events.

In 2020, there was a lot of debate on whether or not COVID-19 ARDS is different from regular pneumonia ARDS^26,27^. Researchers found that ARDS development in COVID-19 patients leads to a higher incidence of acute kidney injury and mortality^28^. It was found that there are lower angiotensin II levels in patients with COVID-19 *and* ARDS compared to patients with milder disease^29^. Thus, physicians may have adjusted their behaviors to monitor renal function more closely in 2021 as they learned more about kidney involvement with ARDS.

For troponin, we observe the opposite relationship– ARDS patients were more closely monitored in 2020 but in 2021 there were no differences between those with and without the disease. A likely explanation is the fact that at the beginning of the pandemic in 2020 there were a lot of uncertainties regarding comorbidities in hospitalized patients. Physicians likely monitored ARDS patients more closely for heart complications just in case. However, by 2021, there is some evidence that cardiac injury is not very associated with ARDS mortality^30^.

For the infection-only labs, we observe the surprising result that *more* missingness is associated with having ARDS. A possible explanation is the fact that there’s really no value in continually monitoring lymphocytes and neutrophils in ARDS patients as it doesn’t generally affect care. Similar to what we see with troponin, in 2020 physicians were generally ordering more tests due to a lack of knowledge of the disease and complications. With ARDS, over time, monitoring other labs likely takes more priority than the infection-only labs.

More missingness in the liver labs was nearly associated with 90-day mortality in 2021 (p = 0.049). Although not significant while controlling for multiple testing, it is still worth investigating with a larger sample size due to the potential implications. COVID-19 patients with ARDS could have potentially better outcomes if their liver labs are monitored more closely. Although not as significant, we also observe a large effect size for more missingness in PT and procalcitonin in 2021 for patients who experienced 90-day mortality. Due to the IPW of propensity scores, we reasoned it might be possible that the significance is tempered by potential confounding by thrombotic events, since PT and procalcitonin are directly involved in monitoring this condition. We found that patients who experienced 90-day mortality had a lower prevalence of thrombotic events. However, we also found that patients with thrombotic events and ARDS tended to be older than patients who only had ARDS. A likely explanation is that older patients with both ARDS and thrombotic events may not have survived long enough to have satisfied the 14-day observation period. This example displays the need to account for potential confounding when assessing relationships between missing data and clinical outcomes so as not to draw false conclusions. The result from the liver labs after accounting for confounding was nearly significant, and could indicate that monitoring liver labs more closely could improve outcomes for COVID-19 ARDS patients. However, this finding requires further investigation.

There are a few limitations to this study. First, our cohort is potentially biased by the fact that long-term patients have to be healthy enough to have survived for 14 days. Fortunately, accounting for confounding by IPW of propensity scores helps prevent us from making any false conclusions that are actually due to selection bias, as we see with the example of PT, procalcitonin, and mortality. However, it may be important to further account for unobserved variables. Second, we only observe patients during 2020 and 2021. Having more recent data to analyze might provide more relevant insights into how missing data associates with ARDS and how missingness predicts 90-day mortality. The sample size for ARDS was also very small (N = 72 in 2020, N = 37 in 2021). When conducting multiple statistical tests and controlling for FDR, it is difficult to find any truly significant results. Thus, the results regarding mortality in ARDS patients should be verified by other studies. For future work, we are interested in incorporating data from other hospitals from the 4CE consortium. We plan to conduct these analyses on 2022 data, and similar to what we did with ARDS, look into other comorbidities more closely such as thrombotic events and neurological prevalence.

## Data Availability

Only aggregate data was shared by sites for this study. All aggregate data in a de-identified fashion can be found and downloaded at www.covidclinical.net.

## Acknowledgments

GSO is supported by NIH grants U24CA210967 and P30ES017885. EG and QL are supported by NIGMS R01GM124111 and NIA RF1AG063481. DLM and JHH are supported by NCATS UL1-TR001878 and U24-DK133700.

## Supplemental Material

### ICD-9 and ICD-10 codes corresponding to thrombotic events

“I74”, “I75”, “I76”, “I21”, “I22”, “I23”, “I26”, “I27”, “Z86”, “I63”, “I67”, “I81”, “I82”,”444”,”445”, “410”,”415”,”V12”,”434”,”437”,”452”,”453”,”D65”, “P60”,”286”,”776”.

### ICD-9 and ICD-10 codes corresponding to neurological events

“R41”,”R27”,”R42”,”G44”,”G03”,”G04”,”G72”,”M60”,”G61”,”G65”,”R43”,”G93”,”F29”,”G40”,”G45”,”G46”,”I 60”,”I61”,”I62”,”I67”,”H54”,”40”,”298”,”307”,”320”,”321”,”322”,”323”,”330”,”331”,”339”,”345”,”348”,”357”,” 359”,”369”,”430”,”431”,”432”

## Notes

### Competing Interest Statement

The authors have declared no competing interest.

### Author Declarations

IRB Approval was obtained at Assistance Publique Hopitaux de Paris, Beth Israel Deaconess Medical Center, Bordeaux University Hospital, Hospital Universitario 12 de Octubre, Massachusetts General Brigham, Northwestern University, Medical Center, University of Freiburg, University of Pittsburgh, VA North Atlantic, VA Southwest, VA Midwest, VA Continental, and VA Pacific. An exempt determination was made by the IRB at University of California Los Angeles, University of Michigan, and University of Pennsylvania.

## References

1. Getzen E, Ungar L, Mowery D, Long Q. Mining for equitable health: assessing the impact of missing data in electronic health records. J Biomed Inform 2023;5:104269

2. Denny J. Mining electronic health records in the genomics era, PLoS Comput. Biol 2012;8:e1002823.

3. Bayley K, Belnap T, Savitz L, Masica A, Shah N, Fleming S. Challenges in using electronic health record data for CER: experience of 4 learning organizations and solutions applied, Med. Care 2013;51:S80–6.

4. Samal L, Dykes P, Greenberg O, Hasan A, Venkatesh A, Volk L, Bates D,. Care coordination gaps due to lack of interoperability in the United States: a qualitative study and literature review, BMC Health Serv. Res 2016;16:143.

5. Shinozaki A. Electronic medical records and machine learning approaches to drug development. Artificial Intelligence in Oncology Drug Discovery and Development 2019.

6. Aerts H, Kalra D, Sáez C, Ramírez-Anguita J, Mayer M, Garcia-Gomez J, Durà-Hernández M, Thienpont G, Coorevits P. Quality of hospital electronic health record (EHR) data based on the international consortium for health outcomes measurement (ICHOM) in heart failure: pilot data quality assessment study, JMIR Med Inform 2021;9:e27842.

7. Argalious M, Dalton J, Sreenivasalu T, O’Hara J, Sessler D. The association of preoperative statin use and acute kidney injury after noncardiac surgery, Anesth. Analg 2022;20 117 (2013) 916–923.

8. Chang C, Deng Y, Jiang X, Long Q. Multiple imputation for analysis of incomplete data in distributed health data networks, Nat. Commun 2020;11:5467.

9. Feldman S, Davlyatov G, Hall A. Toward understanding the value of missing social determinants of health data in care transition planning, Appl. Clin. Inform 2020;11:556–563.

10. Tan A, Getzen E, Hutch M, Strasser Z, Gutiérrez-Sacristán A Le T, Dagliati A, Morris M, Hanauer D, Moal B, Bonzel C, Yuan W, Chiudinelli L, Das P, Zhang H, Aronow B, Avillach P, Brat G, Cai T, Hong C, La Cava W, Shriver E, Shakeri Hossein Abad Z, Tan B, Visweswaran S, Wang X, Weber G, Xia Z, Verdy B. COVID-19 by EHR 4CE Collaborative Group/Consortium, Long Q, Mowery D, Holmes J. Informative missingness: what can we learn from patterns in missing laboratory data in the electronic health record? J Biomed Inform 2023;2:104306.

11. Groenwold, R. Informative missingness in electronic health record systems: the curse of knowing. Diagnostic and Prognostic Research 2020.

12. Singh J, Sato M, Okhuma T. On missingness features in machine learning models for critical care: observational study. MIR Medical Informatics 2021.

13. Butera N, Zeng D, Green Howard A, Gordon-Larsen P, Cai J. A doubly robust method to handle missing multilevel outcome data with application to the China health and Nutrition Survey, Stat. Med 2022;41:769–785.

14. Wu M, Follmann D. Use of summary measures to adjust for informative missingness in repeated measures data with random effects, Biometrics 1999; 55:5–84.

15. Allen A, Collins J, Rathouz P, Selander C, Satten G. Bootstrap calibration of TRANSMIT for informative missingness of parental genotype data, BMC Genet 2003; 4:Suppl 1 S39.

16. Allen A, Rathouz P, Satten G. Informative missingness in genetic association studies: case-parent designs, Am. J. Hum. Genet 2003; 72:671–680.

17. James I, McKinnon E, Gaudieri S, Morahan G, Diabetes Genetics Consortium. Missingness in the T1DGC MHC fine-mapping SNP data: association with HLA genotype and potential influence on genetic association studies, Diabetes Obes. Metab 2009; 11 Suppl 1:101–107.

18. Kujala M, Nevalainen J. A case study of normalization, missing data and variable selection methods in lipidomics, Stat. Med 2015;34:59–73.

19. Lin W, Liu N. Reducing bias of allele frequency estimates by modeling SNP genotype data with informative missingness, Front. Genet 2012;3:107.

20. Lyles R, Allen A, Flanders W, Kupper L, Christensen D. Inference for case-control studies when exposure status is both informatively missing and misclassified, Statistics in Medicine 2006;25:4065–4080.

21. Nicknam Z, Jafari A, Golchin A, Pouya F, Nemati M, Rezaei-Tavirani M, Rasmi Y. Potential therapeutic options for COVID-19: an update on current evidence. European Journal of Medical Research 2022.

22. Liang C, Ogilvie R, Doherty M, Robin Clifford C, Chomistek A, Gately R, Song J, Enger C, Seeger J, Lin N, Wang F. Trends in COVID-19 patient characteristics in a large electronic health record database in the United States: A cohort study. Plos One 2022.

23. Satterfield B, Dikilitas O, Kullo I. Leveraging the electronic health record to address the COVID-19 pandemic. Mayo Clinical Proceedings 2021.

24. Rudolf J, Dighe A, Coley C, Kamis I, Wertheim B, Wright D. Analysis of daily laboratory orders at a large urban academic center: a multifaceted approach to changing test ordering patterns. Am J Clin Pathol 2017; 148:128.

25. Blei D, Ng A, Jordan M. Latent dirichlet allocation, J Mach Learn Res. 2003;3:993–1022.

26. Aslan A, Aslan C, Zolbanin N, Jafari R. Acute respiratory distress syndrome in COVID-19: possible mechanisms and therapeutic management. Pneumonia 2021.

27. Li X, Ma X. Acute respiratory failure in COVID-19: is it “typical” ARDS? Critical Care 2020.

28. Alenezi F, Almeshari M, Mahida R, Bangash M, Thickett D, Patel J. Incidence and risk factors of acute kidney injury in COVID-19 patients with and without acute respiratory distress syndrome (ARDS) during the first wave of COVID-19: a systematic review and Meta-Analysis. Renal Failure 2021.

29. Legrand M, Bell S, Forni L, Joannidis M, Kooyner J, Liu K, Cantaluppi V. Pathophysiology of COVID-19-associated acute kidney injury. Nature Reviews Nephrology 2021.

30. Jayasimhan D, Foster S, Chang C, Hancox R. Cardiac biomarkers in acute respiratory distress syndrome: a systematic review and meta-analysis. Journal of Intensive Care 2021.

